# A case-control study of autoimmune AEFIs following COVID-19 vaccination reported to VAERS

**DOI:** 10.1101/2021.07.06.21260074

**Authors:** Chris von Csefalvay

## Abstract

Autoimmune adverse effects following immunisation (AEFIs) are widely regarded as a chief concern driving vaccine hesitancy. This case-control study seeks to shed light on the true risk of autoimmune AEFIs associated with the COVID-19 vaccine through a case-control analysis of VAERS reports. Reports of autoimmune aetiology were matched with reports of non-autoimmune controls. Statistical analysis reveals that the safety profile of COVID-19 vaccines with regard to autoimmune AEFIs is highly favourable. In particular, neuroautoimmune AEFIs have statistically significant reporting odds ratios below unity (Guillain-Barre syndrome: 0.35, multiple sclerosis: 0.70, transverse myelitis: 0.79), indicating a reduced association of reports of these conditions with the COVID-19 vaccine versus other vaccines. Only three autoimmune aetiologies exceed a ROR of 2.0 and thus present a potential signal. Of these, myasthenia gravis (ROR = 3.90, *p <* 0.001, 95% CI: 2.63-5.80) may be the result of epidemiological confounding factors not sufficiently controlled by matching, as the population most likely to develop myasthenia gravis was strongly prioritised in the COVID-19 vaccine’s initial rollout. Immune thrombocytopaenia (ROR = 26.83, *p <* 0.001, 95% CI: 16.93-42.54) is a clear safety signal, confirming a large number of case reports and studies that indicate a risk of immune thrombocytopaenic events following the COVID-19 vaccine. The lone strong safety signal of immune thrombocytopaenia notwithstanding, this study attests to the safety of the COVID-19 vaccine where autoimmune conditions are concerned. Through quantifying the risk of autoimmune disorders associated with COVID-19 vaccination, this study contributes to a growing body of evidence supporting the safety of such vaccines.

## 1 Introduction

Vaccines are, by their nature, intended to elicit immunogenic effects in the recipient. Where the immune response results in an off-target humoral or cellular immune reaction against the recipient’s own tissue, the clinical picture is typically an autoimmune pathology. The notion of ”vaccine induced autoimmunity”, which altogether overstates how rare and marginal such reactions are, nonetheless numbers among the key controversies in the fight against vaccine-preventable diseases.^[1]^ One must look no further than the story of the ill-fated recombinant hepatitis B vaccine (HBV) for a case study of the devastating effects that may result when the suspicion of inducing autoimmunity is cast upon a vaccine. Despite convincing evidence to the contrary,^[2]^ a number of cases of multiple sclerosis have been attributed to HBV in the late 1990s. Factual evidence for the lack for such an association notwithstanding,^[3]^ the effects in the public consciousness endure. Eurosurveillance reports that in France, where the public discourse about the HBV-MS association was perhaps the most intense, hepatitis B vaccination uptake was consistently below 50% in all segments of the population^[4]^ – a long way from the recommendation of universal hepatitis B vaccination.

The endeavour to reach collective immunity to COVID-19 by way of vaccination is, in this sense, no less susceptible to the danger of vaccine hesitancy.^[5]^ Qualitative studies attest that autoimmunity is among the key concerns of those refusing vaccination, even among allied health professionals.^[6]^ This is exacerbated by the dramatic rise in misinformation about the safety of the COVID-19 vaccines,^[7]^ especially via social media platforms like TikTok^[8]^ and Twitter.^[9,10]^ In addition, two of the three vaccines approved in the United States for COVID-19 prophylaxis at the time of writing use mRNA technology to create the SARS-CoV-2 Spike protein antigen in the recipient’s body. The two mRNA vaccines, widely known as the Pfizer/BioNTech and the Moderna vaccines (BNT162b2, Pfizer, Inc., Collegeville, PA, USA and mRNA-1273/elasomeran, ModernaTX, Inc., Cambridge, MA, USA, respectively), are the first prophylactic vaccines to be approved by the Food and Drug Administration that rely on messenger RNA. Both the ’novelty factor’, the emergency circumstances of authorisation and misunderstandings about the mechanism of action have contributed to complex public sentiment on the matter.

The purpose of this paper is to provide a degree of resolution to that complexity by examining early information on autoimmune aetiologies submitted to the Vaccine Adverse Event Reporting System (VAERS) maintained jointly by the CDC and the FDA. At the time of writing, over 330m doses of the COVID-19 vaccines have been administered in the United States, with a little under half of the population being fully vaccinated. The speed and effectiveness of this ambitious vaccination project is no doubt historical, but whether it reaches the principal aim of attaining or approaching herd immunity depends largely on the next few months. By quantitatively assessing the true risk of autoimmune adverse effects on the basis of passive reporting to VAERS, this paper contributes to dispelling concerns about off-target immunogenic effects and autoimmune presentations resulting from the COVID-19 vaccines.

## 2 Materials and methods

### 2.1 Data sources

Reporting data was obtained from VAERS via vaers.hhs.gov on 04 July 2021, with data including reports up to 02 July 2021. In order to ensure a sufficient number of controls, data was also obtained going back to 01 January 1995, inclusive.

Maintained jointly by the CDC and the FDA, VAERS is a passive reporting system for potentially vaccine-related adverse events.^[11]^ As a passive reporting system, it relies on healthcare professionals, administrators and recipients of vaccines to submit information about potential adverse events following immunisations (AEFIs). Reports may be made anonymously, and there is no verification of reports against other objective sources, such as death certificates for alleged fatal AEFIs or against hospital EMRs for claimed admissions. Nor is there an established system to deduplicate reports that may have been submitted by multiple parties to the same event, potentially without each other’s knowledge.^[12]^ This results in a significant potential for overreporting of AEFIs to VAERS.

At the same time, there is no general obligation to report most AEFIs to VAERS. An exception under this are circumstances covered by the National Childhood Vaccine Injury Act of 1986 (42 U.S.C. *§*300aa-1 to 300aa-34). Each of the Emergency Use Authorizations under which the three COVID-19 vaccines were authorised in the United States mandate reporting of certain AEFIs, specifically

- any and all administration errors,
- serious adverse effects (AEFIs that result in death, life-threatening illness, inpatient hospitalisation or prolongation of pre-existing hospitalisation, persistent disability or which are determined to be serious in medical judgment),
- instances of Multisystem Inflammatory Syndrome, and
- cases of COVID-19 necessitating hospitalisation or resulting in death.

As a passive reporting system, even with this increased degree of mandatory reporting for COVID-19 vaccines, some AEFIs will go unreported. The very extent of this underreporting is hard to ascertain.^[13]^ Therefore, the study design was devised so as to mitigate the effects of under- and overreporting as long as these are uniform across vaccines.

Data loading and management was performed using Python 3.7.5 and `pandas` 1.3.0.^[14]^

### 2.2 Identifying cases

Cases were obtained from VAERS reports received between 1 January 1995 and 2 July 2021, inclusive, in respect of vaccinations that took place within the United States. A report was considered a case if at least one of the entries in the reported SYMPTOM fields belonged to the set of relevant diagnoses. A diagnosis was considered relevant if its MedDRA code fell within the MedDRA High Level Group Terms (HLGT) for autoimmune disorders (10003816) or immune disorders NEC (10027665), but specifically exclude sarcoidoses (10039487), amyloidoses (10002023), blood isoimmune reactions (10023053) and transplant rejection (10052779). In addition, six diagnoses were explicitly excluded: ARDS (10001052) and Systemic Immune Response Syndrome (SIRS) (10051379), which are insufficiently specific to an autoimmune cause in the COVID-19 context, skin sensitisation (10040785) as it is principally of an allergic rather than autoimmune etiology, shoulder injury related to vaccine administration (10081038) as it permits a wide non-autoimmune etiology, and the general categories of ”adverse event following immunisation” (10069520) and ”vaccination complication” (10046861), which were insufficiently specific. This yielded the following hierarchy (included diagnoses in bold type):

- Immune system disorders (10021428)
  - Autoimmune disorders (10003816)
    * **Autoimmune disorders NEC (10027657)**
    * **Blood autoimmune disorders (10003817)**
    * **Endocrine autoimmune disorders (10003818)**
    * **Hepatic autoimmune disorders (10003820)**
    * **Lupus erythematosus and associated conditions (10025136)**
    * **Muscular autoimmune disorders (10003821)**
    * **Nervous system autoimmune disorders (10074484)**
    * **Rheumatoid arthritis and associated conditions (10039075)**
    * **Scleroderma and associated disorders (10039711)**
    * **Skin autoimmune disorders NEC (10052738)**
  - Immune disorders NEC (10027665)
    * Acute and chronic sarcoidosis (10039487)
    * Amyloidoses (10002023)
    * **Autoinflammatory diseases (10073080)**
    * *∗*Blood isoimmune reactions (10023053)
    * **Immune and associated conditions NEC (10027682)**, except:
      · ARDS (10001052),
      · Adverse event following immunisation (10069520),
      · Shoulder injury related to vaccine administration (10081038),
      · Vaccination complication (10046861),
      · Skin sensitisation (10040785), and
      · Systemic Immune Response Syndrome (SIRS) (10051379).
    * Transplant rejection (10052779)
    * **Vasculitides (10052779)**

Altogether, this yielded 9,287 cases, of whom 2,318 (24.96%) have been exposed. A detailed cross-tabulation of the results is found in Table 1.

**Table 1:**
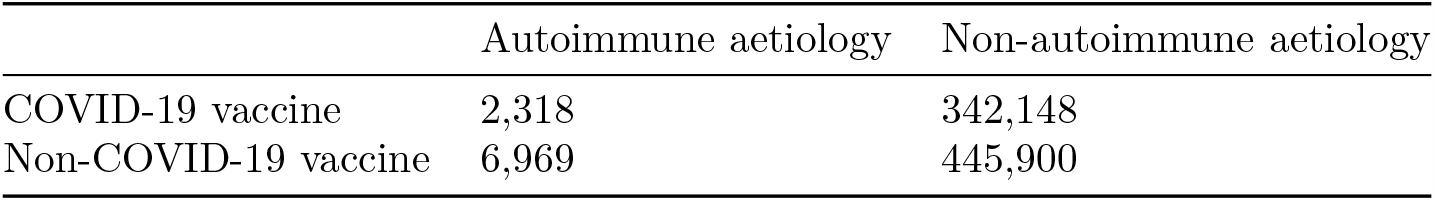
Cross-tabulation of cases and controls (columns) by exposure status (rows).

### 2.3 Identifying controls

For the purposes of this analysis, we consider reports a potential control if none of the reported symptoms fall within the autoimmune aetiologies referred to in Subsection 2.2. Potential controls were, like cases, obtained from VAERS reports between 1 January 1995 and 2 July 2021. This yielded a pool of 788,048 potential controls.

### 2.4 Identifying exposure

An exposure was defined as the listing of a COVID-19 vaccine in a report. Notably, if the report listed the COVID-19 vaccine in the patient’s prior medical history or medications field, but not in the field of vaccines in respect of which the report is being filed, the report was not considered to be part of the exposed class.

A COVID-19 vaccine, for the purposes of this analysis, was defined as any of the approved COVID-19 vaccines in the United States, i.e. any vaccine with a VAERS `VAX_TYPE` of `COVID19`. This included VAERS reports that specified the `VAX_TYPE` as a COVID-19 vaccine, but did not identify the specific manufacturer, recording the vaccine as `COVID19 (UNKNOWN)`.

### 2.5 Case-control matching

Cases and controls were matched using `pymatch` 0.3.4. For each case, three controls were selected randomly from all potential controls that match the case’s gender (male, female or unknown/unspecified) and the case’s age band (*<*18, 18-25, 26-40, 41-55, 56-70 and *>*70). Separability was assessed by fitting 100 sequential models on balanced samples, with an average return of 59.3% indicating some mild separability in the data. In particular, as is common with age-banded data, propensities exhibited a hexamodal distribution that reflected the number of age bands, with separability emerging most clearly towards the extremes. The pre-match propensity score distribution is presented as Figure 1.

**Figure 1:**
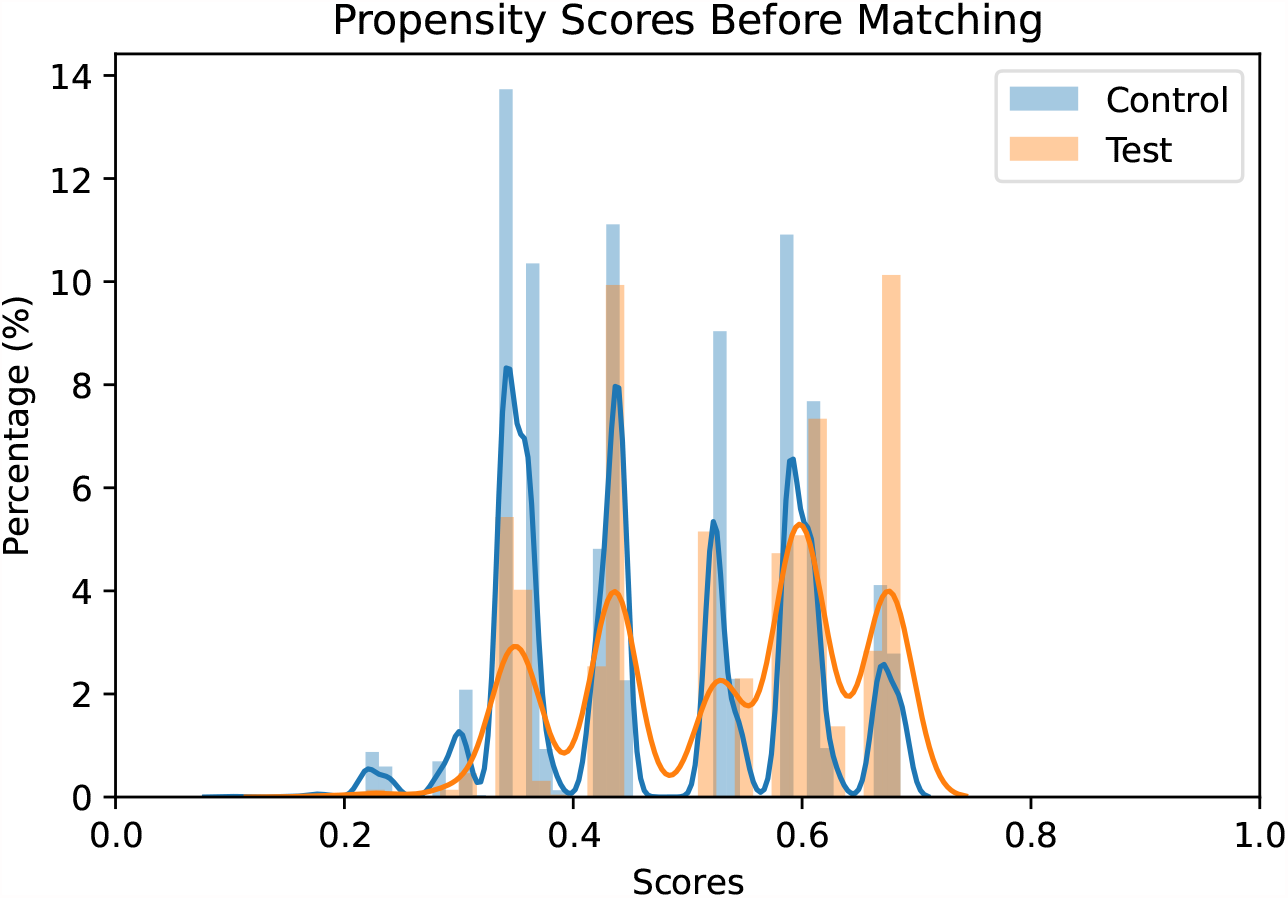
Propensity scores of the case and control populations before matching.

### 2.6 Statistical analysis

Reporting odds ratios (ROR) were calculated using Fisher’s exact test from `scipy` 1.7.0 for all conditions in the list of autoimmune aetiologies in Subsection 2.2 where at least 50 cases could be identified in the exposed population.

The ROR was calculated against all symptoms except certain purely diagnostic entities (e.g. mere mentions of a test), procedures, ongoing care (e.g. pace-maker maintenance) and normal test results. The statistical significance was determined by calculating two-sided *p* values using Fisher’s exact test, under the null hypothesis that reported cases and reported controls would have the same likelihood of exposure to a COVID-19 vaccine.

## 3 Results

As Figure 2 shows, results were statistically significant for all but two AEFIs: unspecified vasculitis (MedDRA 10047115) and unspecified autoimmune disorder (MedDRA 10061664). The RORs are particularly low for neuroautoimmune conditions (Guillain-Barre syndrome, optic neuritis, multiple sclerosis and transverse myelitis). This confirms previous findings as to the safety of COVID-19 vaccines with respect to neuroautoimmune AEFIs.^[12]^

**Figure 2:**
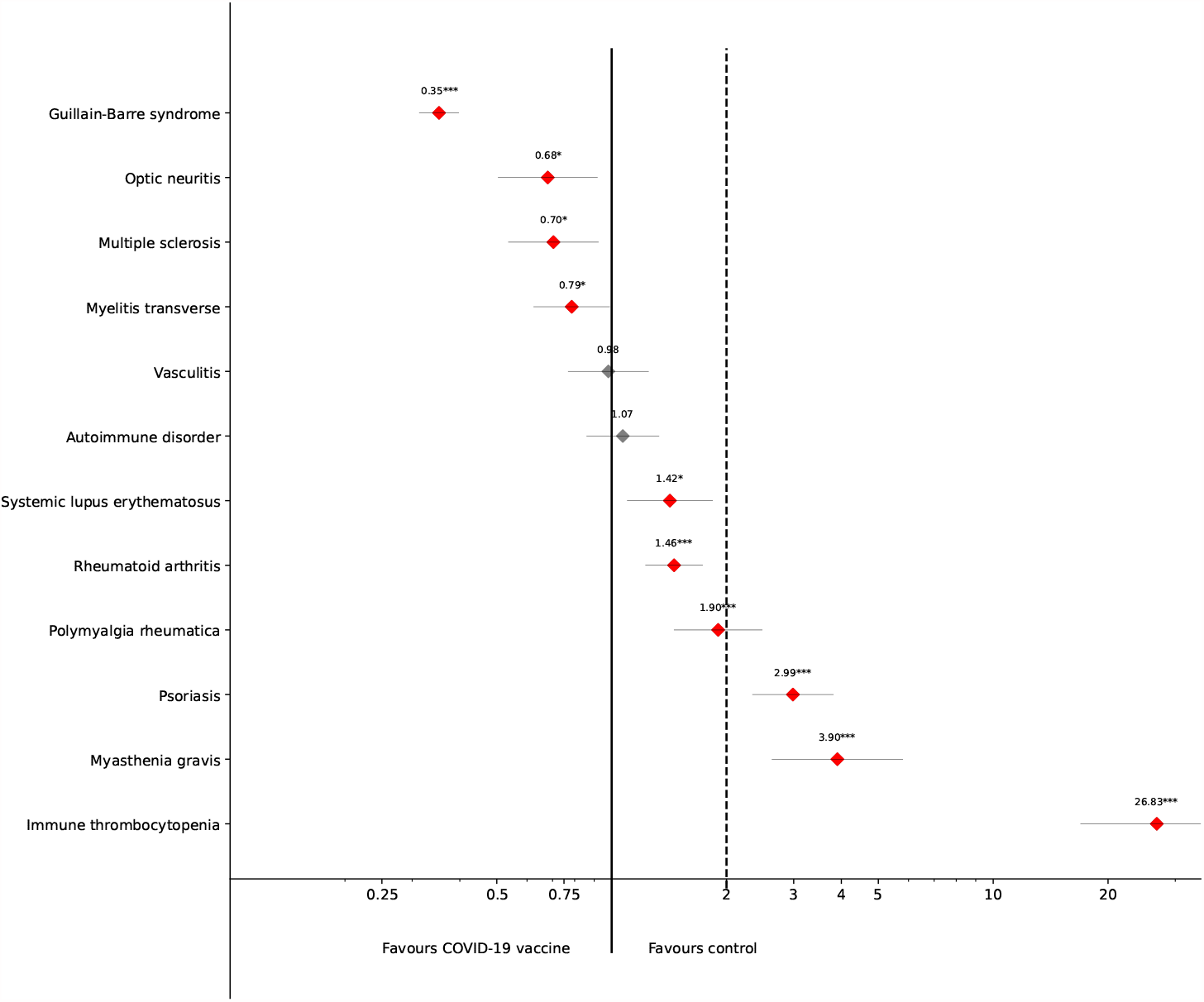
Forest plot of autoimmune AEFIs with at least 50 cases in the treatment group by Reporting Odds Ratio. Red markers indicate results that are statistically significant at least at *p <* 0.05.

Only three specific autoimmune aetiologies exceed the reporting odds ratio of 2.0, which is widely considered the minimum for signal generation in passive pharmacovigilance. Psoriasis (ROR = 2.99, *p <* 0.001, 95% CI: 2.34-3.82) and myasthenia gravis (ROR = 3.90, *p <* 0.001, 95% CI: 2.63-5.80) were observed with a somewhat elevated ROR vis-a-vis other vaccines and other potential AEFIs. In this age- and gender-matched analysis, the most significant result that favoured controls pertained no doubt to immune thrombocytopaenia. As Figure 2 indicates, the ROR for immune thrombocytopaenia was 26.83 (*p <* 0.001, 95% CI: 16.93-42.54) - in other words, a patient who presented with immune thrombocytopaenia after a vaccination is approx 27 times more likely to have received a COVID-19 vaccine than a non-COVID-19 vaccine. Nevertheless, this must be understood in the context of the COVID-19 vaccine itself, and in particular in the context of the confounding effects of age and age-related factors.

## 4 Discussion

### 4.1 Neuroautoimmune AEFIs

This study confirms previous findings as to the safety of the COVID-19 vaccine with regard to neuroautoimmune disorders.^[12]^ In particular, with respect to Guillain-Barre syndrome, patients were three times as likely to have received a non-COVID-19 vaccine than a COVID-19 vaccine. The absence of a neuroautoimmune safety signal may be usefully emphasised in reassuring the population about the COVID-19 vaccines, especially considering the prominent position such side effects occupy in the landscape of concerns about vaccines that drive vaccine hesitancy.^[6]^ Neuroautoimmune disorders are serious, potentially fatal and in many cases permanently disabling. For this reason, such an apprehensive reaction is hardly unintelligible. The findings of this study further contribute to the growing evidence base that underlines the safety of COVID-19 vaccines with regard to neuroautoimmunity.

### 4.2 Myasthenia gravis

Myasthenia gravis is an autoimmune process in which autoantibodies interfere with neuromuscular signalling at the neuromuscular junction, specifically with the nicotinic acetylcholine receptor (nAChR).^[15]^ Other than a case study of myasthenia gravis crisis following COVID-19 vaccination,^[16]^ there has been generally no evidence of a myasthenia gravis-related safety signal. In fact, the clinical consensus is that the COVID-19 vaccine is safe for patients with myasthenia gravis, and indeed recommended as myasthenia gravis might correlate to a worse clinical outcome with COVID-19.^[17,18]^

The slightly elevated ROR must be understood in the context of the COVID-19 vaccination programme. First, the incidence of myasthenia gravis increases with age.^[19,20]^ While the present study controls for age as a confounding factor to some extent by way of case-control matching, the effect of such tools on obviating confounders must not be overestimated.^[21]^ Case-control matching may reduce confounders, but certain higher order effects may remain resistant to this. In particular, the COVID-19 vaccine was initially targeted at patients with pre-existing conditions who might develop worse clinical outcomes if experiencing COVID-19 infection. For this reason, it is quite likely that the cohort of those who have received the COVID-19 vaccine would have had a higher prevalence of chronic health conditions along with a higher average age. Indeed, in many parts of the United States, vaccinations were primarily available only for those above a certain age and/or suffering from chronic health conditions (’high risk patients’). Given the overall worse outcomes of COVID-19 among the population with neuromuscular disorders overall,https://doi.org/10.1196/annals.1254.053 it is therefore likely that such patients were overrepresented.

An additional factor is the not infrequent paraneoplastic etiology of myasthenia gravis. Lambert-Eaton myasthenic syndrome (LEMS) is a paraneoplastic syndrome associated most often with small cell lung cancer (SCLC).^[22]^ Although Lambert-Eaton myasthenic syndrome has a MedDRA code of its own (10067685), it is not inconceivable that a proportion of the myasthenia gravis cases reported are of a paraneoplastic aetiology. Patients with malignancies, especially those undergoing cytotoxic and/or immunosuppressive chemotherapy, have been among the first priority groups of the COVID-19 vaccination effort throughout the United States. In addition, patients with previous episodes of myasthenia gravis are frequently advised to avoid live vaccines,^[23]^ and evidence to the safety of most vaccines in myasthenia gravis notwithstanding, many physicians recommend against other vaccinations as well. ^[24]^ Overall, these factors could plausibly have combined to increase the apparent frequency of reported myasthenia gravis episodes (which may be first or subsequent) to VAERS following COVID-19 vaccination. In the absence of a solid biological causal mechanism, and given the relatively low ROR, the evidence continues to support the professional consensus that patients with myasthenia gravis may safely receive the COVID-19 vaccine.^[18]^ The risk of developing myasthenia gravis is, in view of potential confounding factors, unlikely to be strongly associated with the COVID-19 vaccine.

### 4.3 Immune thrombocytopaenia

Immune thrombocytopaenia (until recently also known as immune thrombocytopaenic purpura, ITP) covers conditions of isolated thrombocytopaenia in the absence of bone marrow dysplasia.^[25]^ Like myasthenia gravis, immune thrombocytopaenia is associated with factors that predispose towards a higher priority for vaccination. Immune thrombocytopaenia typically distributes along age in a bimodal manner, with peaks in early childhood and above the age of 60, and approx. 12% of patients with immune thrombocytopaenia in a recent French study exhibited malignancies or myelodysplasia.^[26]^ As such, the presence of uncontrolled confounding factors in this case is plausible to some extent.

Nevertheless, the very high relative reporting ratio of 26.83 cannot be discounted purely by reference to confounding factors. Cases of post-COVID-19 vaccine immune thrombocytopaenia have been widely reported in the literature,^[27–30]^ across a range of vaccines, including the Moderna vaccine as well as vaccines like ChAdOx1 (widely known as the Oxford-AstraZeneca vaccine). While no causal mechanism has been convincingly identified for immune thrombocytopaenia following COVID-19 vaccination, molecular mimicry appears to be a plausible potential mechanism.^[31]^

The association of the Oxford-AstraZeneca vaccine with immune thrombocytopaenia has been widely reported upon.^[32]^ This may induce a degree of misattribution bias and even misdiagnosis (since in approx. a third of cases diagnosed as immune thrombocytopaenia, no autoantibodies are expressly identified). On the other hand, the high ROR incontrovertbly suggests a potential safety signal. Welsh *et al*. convincingly demonstrate that despite this potential signal, the number of reports of immune thrombocytopaenia in view of the number of vaccines administered does not exceed expectations.^[33]^ Thus, while the high ROR of immune thrombocytopaenia is no doubt putting COVID-19 vaccination on the map as a potentially associated exposure, it does not on its own suggests an association. Out of an abundance of caution, further research and monitoring of immune thrombocytopaenic episodes following COVID-19 vaccination may nevertheless be advisable.

### 4.4 Limitations

As any study that relies on passive reporting, data in this study must be considered in the wider context of limitations to passive reporting in general and VAERS in particular. First, such analyses rely on what is reported, suffering not only from under- and overreporting at the same time but also the phenomenon known as differential reporting.^[34]^ At the heart of differential reporting is the cognitive bias that events seem to be related if they are temporally closely related and, vice versa, they are seen to be independent if they are further apart. Since autoimmune disorders in particular often develop over a longer period of time or may be latently present (from a biochemical perspective, i.e. presence of autoantibodies) well before symptoms are felt and noticed, some cases may evade association, and thus also evade reporting. Equally, many autoimmune disorders have a relatively long time to diagnosis due to the often nonspecific symptoms. Finally, VAERS accepts reports from lay reporters (e.g. patients and parents) whose attribution may not necessarily be in line with commonly accepted diagnostic categories. While the study design has been drafted to reduce the risk of these factors through matching and through comparison with other reported symptoms, its results must be seen with the limitations of VAERS firmly kept in mind.

## 5 Conclusion

Case-control studies from passive reporting may contribute to our understanding of the wider adversome of COVID-19 vaccines. While passive reporting carries with itself inherent sources of bias and both under- and overreporting, such sources are valuable for generating early signals ”in the wild”. This early analysis contributes to our understanding of the COVID-19 vaccine’s adverse effects profile by focusing on autoimmune AEFIs in particular, concluding that for most potential AEFIs, an association with a COVID-19 vaccine is much less likely than an association with a non-COVID-19 vaccine.

Only a small minority of AEFIs appear to show a potential safety signal. Of these, the safety signals of psoriasis and myasthenia gravis are significantly less pronounced. In particular, the potential safety signal of myasthenia gravis is to a great extent explained – and thus, contradicted – by the strong association of myasthenia gravis with factors such as age, which is reported to VAERS in a structured manner, and neoplastic comorbidity, which is not.

This study underlines to a great extent the widely shared professional understanding that the COVID-19 vaccine is not liable to induce autoimmune disorders or exacerbate existing autoimmune pathologies.^[35–37]^ In this sense, it is a ’negative study’^[38]^ – after all, rather than highlighting a new source of risk, the findings above primarily reflect the safety of the COVID-19 vaccine in the context of autoimmune AEFIs. It is, however, also a positive finding in the sense that it may serve to bolster the case in favour of vaccination among populations apprehensive of vaccinations due to the possibility of autoimmune side effects or among patients already affected by an autoimmune disorder.

The single AEFI of concern is immune thrombocytopaenia (see Subsection 4.3), which has already been addressed both as case studies^[27–30]^ and as in-depth epidemiological studies.^[33]^ In that sense, the safety signal generated by this study has already been picked up and analysed in detail. Further research is certainly appropriate to monitor the expanding wealth of data on COVID-19 vaccination. Nevertheless, the evidence from this study, based on over six months of vaccinations and over 300 million individual vaccines, certainly confirms the consensus that COVID-19 vaccines are not significantly associated with any unknown autoimmune risk factors. This result may be usefully integrated not only into clinical care but also the wider discourse when responding to patient concerns about potential off-target immunogenic effects of COVID-19 vaccines.

## Data Availability

VAERS data can be obtained from vaers.hhs.gov.
Supplementary material S1, a CSV (comma-separated values) version of the data underlying Figure 2, is available as part of DOI 10.5281/zenodo.5075754.
All code and materials are available on Github under https://github.com/chrisvoncsefalvay/covid-19-autoimmune-aefis.

https://vaers.hhs.gov/data.html

https://github.com/chrisvoncsefalvay/covid-19-autoimmune-aefis

## Supplementary material

Supplementary material S1, a CSV (comma-separated values) version of the data underlying Figure 2, is available as part of DOI 10.5281/zenodo.5075754.

## Funding

This research was funded by Starschema Inc. under its intramural research funding programme.

## Data availability

VAERS reporting data is available from the CDC’s website at https://vaers.hhs.gov. All code and scripts supporting this manuscript are deposited at https://github.com/chrisvoncsefalvay/covid-19-autoimmune-aefis and are made available under the DOI 10.5281/zenodo.5075754.

## Conflicts of interest

CvC is a consultant to a company that may be affected by the research reported in this paper. The funders had no role in the design of the study; in the collection, analysis, or interpretation of data; in the writing of the manuscript, or in the decision to publish the results.

## Notes

### Author Declarations

No IRB approval was sought as the study is a pure retrospective analysis over data reported to VAERS and thus publicly available.

## References

[1] Frank DeStefano, Heather Monk Bodenstab, and Paul A Offit. Principal Controversies in Vaccine Safety in the United States. Clinical Infectious Diseases, 69(4):726–731, 02 2019. ISSN 1058-4838. doi: 10.1093/cid/ciz135. URL https://doi.org/10.1093/cid/ciz135.

[2] Alberto Ascherio, Shumin M. Zhang Miguel, A. Hernán, Michael J. Olek, Paul M. Coplan, Kimberly Brodovicz, and Alexander M. Walker. Hepatitis B Vaccination and the Risk of Multiple Sclerosis. New England Journal of Medicine, 344(5):327–332, 2001. doi: 10.1056/NEJM200102013440502. URL https://doi.org/10.1056/NEJM200102013440502. PMID: 11172163.

[3] Frank DeStefano, Thomas Verstraeten, and Robert T Chen. Hepatitis B vaccine and risk of multiple sclerosis. Expert Review of Vaccines, 1(4):461– 466, dec 2002. doi: 10.1586/14760584.1.4.461. URL https://doi.org/10.1586%2F14760584.1.4.461.

[4] Dominique Rey, Lisa Fressard, Sébastien Cortaredona, Aurélie Bocquier, Arnaud Gautier, Patrick Peretti-Watel, and Pierre Verger. Vaccine hesitancy in the French population in 2016, and its association with vaccine uptake and perceived vaccine risk–benefit balance. Eurosurveillance, 23(17), apr 2018. doi: 10.2807/1560-7917.es.2018.23.17.17-00816. URL https://doi.org/10.2807%2F1560-7917.es.2018.23.17.17-00816.

[5] Amiel A Dror, Netanel Eisenbach, Shahar Taiber, Nicole G Morozov, Matti Mizrachi, Asaf Zigron, Samer Srouji, and Eyal Sela. Vaccine hesitancy: the next challenge in the fight against COVID-19. European Journal of Epidemiology, 35(8):775–779, 2020.

[6] Sarah D Berry, Kimberly S Johnson, Lonnita Myles, Laurie Herndon, Ana Montoya, Shekinah Fashaw, and David Gifford. Lessons learned from front-line skilled nursing facility staff regarding COVID-19 vaccine hesitancy. Journal of the American Geriatrics Society, 2021.

[7] Md Saiful Islam, Abu-Hena Mostofa Kamal, Alamgir Kabir, Dorothy L Southern, Sazzad Hossain Khan, SM Murshid Hasan, Tonmoy Sarkar, Shayla Sharmin, Shiuli Das, Tuhin Roy, et al. Covid-19 vaccine rumors and conspiracy theories: The need for cognitive inoculation against misinformation to improve vaccine adherence. PloS one, 16(5):e0251605, 2021.

[8] Corey H Basch, Zoe Meleo-Erwin, Joseph Fera, Christie Jaime, and Charles E Basch. A global pandemic in the time of viral memes: Covid-19 vaccine misinformation and disinformation on tiktok. Human Vaccines & Immunotherapeutics, pages 1–5, 2021.

[9] Matthew D Kearney, Shawn C Chiang, and Philip M Massey. The Twitter origins and evolution of the COVID-19 “plandemic” conspiracy theory. Harvard Kennedy School Misinformation Review, 1(3), 2020.

[10] Chayakrit Krittanawong, Bharat Narasimhan, Hafeez Ul Hassan Virk, Harish Narasimhan, Joshua Hahn, Zhen Wang, and WH Wilson Tang. Misinformation dissemination in twitter in the COVID-19 era. The American Journal of Medicine, 133(12):1367, 2020.

[11] Robert T Chen, Suresh C Rastogi, John R Mullen, Scott W Hayes, Stephen L Cochi, Jerome A Donlon, and Steven G Wassilak. The Vaccine Adverse Event Reporting System (VAERS). Vaccine, 12(6):542–550, 1994.

[12] Chris von Csefalvay. VAERS data reveals no increased risk of neuroautoimmune adverse events from COVID-19 vaccines. medRxiv, 2021. doi: 10.1101/2021.06.13.21258851.

[13] James A. Singleton, Jenifer C. Lloyd, Gina T. Mootrey, Marcel E. Salive, and Robert T. Chen. An overview of the vaccine adverse event reporting system (VAERS) as a surveillance system. Vaccine, 17(22):2908–2917, 1999. ISSN 0264-410X. doi: https://doi.org/10.1016/S0264-410X(99)00132-2. URL https://www.sciencedirect.com/science/article/pii/S0264410X99001322.

[14] Wes McKinney et al. pandas: a foundational python library for data analysis and statistics. Python for High Performance and Scientific Computing, 14(9):1–9, 2011.

[15] Bianca M Conti-Fine, Monica Milani, Henry J Kaminski, et al. Myasthenia gravis: past, present, and future. The Journal of Clinical Investigation, 116 (11):2843–2854, 2006.

[16] Ariana R Tagliaferri, Spandana Narvaneni, Mohd. Hazem Azzam, and William Grist. A case of COVID-19 vaccine causing a myasthenia gravis crisis. Cureus, jun 2021. doi: 10.7759/cureus.15581. URL https://doi.org/10.7759%2Fcureus.15581.

[17] Saiju Jacob, Srikanth Muppidi, Amanda Guidon, Jeffrey Guptill, Michael Hehir, James F Howard, Isabel Illa, Renato Mantegazza, Hiroyuki Murai, Kimiaki Utsugisawa, et al. Guidance for the management of myasthenia gravis (MG) and Lambert-Eaton myasthenic syndrome (LEMS) during the COVID-19 pandemic. Journal of the Neurological Sciences, 412, 2020.

[18] Sasha A Zîvković, Gregory Gruener, Pushpa Narayanaswami, AANEM Quality, and Patient Safety Committee. Doctor—Should I get the COVID-19 vaccine? Infection and immunization in individuals with neuromuscular disorders. Muscle & Nerve, 63(3):294–303, 2021.

[19] Lawrence H Phillips. The epidemiology of myasthenia gravis. Annals of the New York Academy of Sciences, 998(1):407–412, 2003. doi: https://doi.org/10.1196/annals.1254.053. URL https://nyaspubs.onlinelibrary.wiley.com/doi/abs/10.1196/annals.1254.053.

[20] Lawrence H Phillips, James C Torner, Michael S Anderson, and Gary M Cox. The epidemiology of myasthenia gravis in central and western virginia. Neurology, 42(10):1888–1888, 1992.

[21] Mohammad Ali Mansournia, Nicholas Patrick Jewell, and Sander Greenland. Case–control matching: effects, misconceptions, and recommendations. European Journal of Epidemiology, 33(1):5–14, 2018.

[22] J. O’neill, NMF Murray, and J Newsom-Davis. The Lambert-Eaton myasthenic syndrome: a review of 50 cases. Brain, 111(3):577–596, 1988.

[23] Nils Erik Gilhus, Fredrik Romi, Yu Hong, and Geir Olve Skeie. Myasthenia gravis and infectious disease. Journal of Neurology, 265(6):1251–1258, 2018.

[24] Lorne Zinman, Jessica Thoma, Jeffrey C Kwong, Alex Kopp, Therese A Stukel, and David N Juurlink. Safety of influenza vaccination in patients with myasthenia gravis: a population-based study. Muscle & Nerve: Official Journal of the American Association of Electrodiagnostic Medicine, 40 (6):947–951, 2009.

[25] Gaurav Kistangari and Keith R McCrae. Immune thrombocytopenia. Hematology/Oncology Clinics, 27(3):495–520, 2013.

[26] Guillaume Moulis, Aurore Palmaro, Jean-Louis Montastruc, Bertrand Godeau, Maryse Lapeyre-Mestre, and Laurent Sailler. Epidemiology of incident immune thrombocytopenia: a nationwide population-based study in France. Blood, The Journal of the American Society of Hematology, 124 (22):3308–3315, 2014.

[27] Cheng Xie, Laura Vincent, Andrew Chadwick, and Heiko Peschl. Covid-19 vaccine induced prothrombotic immune thrombocytopenia. European Heart Journal, 2021.

[28] Omar Tarawneh and Husam Tarawneh. Immune thrombocytopenia in a 22-year-old post Covid-19 vaccine. American Journal of Hematology, 2021.

[29] Andreas Tiede, Ulrich J Sachs, Andreas Czwalinna, Sonja Werwitzke, Rolf Bikker, Joachim K Krauss, Frank Donnerstag, Karin Weißenborn, Günter Höglinger, Benjamin Maasoumy, et al. Prothrombotic immune thrombocytopenia after COVID-19 vaccine. Blood, 2021.

[30] M Pai, A Grill, N Ivers, A Maltsev, K Miller, F Razak, M Schull, B Schwartz, NM Stall, R Steiner, et al. Vaccine-induced prothrombotic immune thrombocytopenia (VIPIT) following AstraZeneca COVID-19 vaccination. Science briefs of the Ontario COVID-19 science advisory table, 1 (17), 2021.

[31] Vrushali Saudagar, Satish Patil, Shaun Goh, and Sohil Pothiawala. Vigilance regarding immune thrombocytopenic purpura after covid-19 vaccine. Irish Journal of Medical Science (1971-), pages 1–2, 2021.

[32] Allyson M Pishko, James B Bussel, and Douglas B Cines. Covid-19 vaccination and immune thrombocytopenia. Nature Medicine, pages 1–2, 2021.

[33] Kerry J. Welsh, Jane Baumblatt, Wambui Chege, Ravi Goud, and Narayan Nair. Thrombocytopenia including immune thrombocytopenia after receipt of mRNA COVID-19 vaccines reported to the Vaccine Adverse Event Reporting System (VAERS). Vaccine, 39(25):3329–3332, 2021. ISSN 0264-410X. doi: https://doi.org/10.1016/j.vaccine.2021.04.054. URL https://www.sciencedirect.com/science/article/pii/S0264410X21005247.

[34] Steven Rosenthal and Robert Chen. The reporting sensitivities of two passive surveillance systems for vaccine adverse events. American Journal of Public Health., 85(12):1706–1709, 1995.

[35] Mehmet Soy, Gökhan Keser, Pamir Atagunduz, Melek Yal·in Mutlu, Alper Gunduz, Gizem Koybaşi, and Cemal Bes. A practical approach for vaccinations including COVID-19 in autoimmune/autoinflammatory rheumatic diseases: a non-systematic review. Clinical Rheumatology, pages 1–13, 2021.

[36] Zahid Ali, Musharraf Sarwar, Sheraz Ansar, Usman Ayub Awan, Haroon Ahmed, Nauman Aftab, and Muhammad Sohail Afzal. Covid-19 vaccination hesitancy in patients with autoimmune diseases: A mystery that needs an immediate solution! Journal of Medical Virology, 2021.

[37] Victoria Furer, Christien Rondaan, Nancy Agmon-Levin, Sander Van Assen, Marc Bijl, Meliha Crnkic Kapetanovic, Annette De Thurah, Ulf Mueller-Ladner, Daphna Paran, Karen Schreiber, et al. Point of view on the vaccination against COVID-19 in patients with autoimmune inflammatory rheumatic diseases. RMD open, 7(1):e001594, 2021.

[38] Bruce G. Gellin and William Schaffner. The risk of vaccination — the importance of “negative” studies. New England Journal of Medicine, 344 (5):372–373, 2001. doi: 10.1056/NEJM200102013440509. URL https://doi.org/10.1056/NEJM200102013440509. PMID: 11172170.

